# Comparative Analysis of Inflammatory Response in Surgical Wound Drainage Fluid in Scoliosis Surgery: A Study of Neuromuscular vs. Idiopathic Patients

**DOI:** 10.1101/2025.04.10.25325589

**Authors:** Meredith J. Crane, Robin L. McKinney, Alexander R. D. Jordon, Craig P. Eberson, Amanda M. Jamieson

## Abstract

This study examines immune and inflammatory responses in draining wound fluid over the course of the early stages of wound healing in patients recovering from spinal fusion surgery. The inflammatory phase of wound healing is essential for setting the stage for successful tissue repair and preventing chronic or poorly healing wounds. Scoliosis can be idiopathic or occur secondary to neuromuscular disorders, which are known to be associated with poor wound healing outcomes. We hypothesized that neuromuscular scoliosis patients would exhibit differences in inflammatory wound healing markers compared to idiopathic scoliosis patients. Comparison of the cellular and cytokine contents of draining wound fluid revealed that several inflammatory cytokines were elevated in the neuromuscular scoliosis patient group compared to idiopathic, whereas the leukocyte contents were the same between groups. This study shows that draining wound fluid is a good source of cellular and soluble biomarkers for acute wound healing and can be used to determine changes in individuals at risk for wound healing complications.

## Introduction

Wound healing is a complex process that relies on coordinated stages to progress from injury to repair. The initial phase of wound healing is inflammatory. Neutrophil recruitment occurs within hours of injury in response to platelet-derived chemotactic factors, edema, and cellular and microbial debris. This drives a cascade of inflammation characterized by cytokine release and innate leukocyte recruitment. Controlled early inflammation is necessary for the removal of cell and tissue debris and to combat infection before the wound transitions to the reparative stages of healing. Disruptions to the inflammatory phase can lead to pathological wound healing.^1–6^ Impaired inflammation increases susceptibility to infection and delays the progression to repair, while over-exuberant inflammation can lead to chronic wounds.^7–17^ The work presented here assessed the impact of neuromuscular conditions on the inflammatory response to surgical injury in a cohort of scoliosis patients. Understanding how wound healing is disrupted in certain cohorts of patients could lead to better treatments.

Scoliosis is a three-dimensional spinal deformity that can lead to secondary health effects such as back pain, pulmonary dysfunction, and disfigurement.^18–20^ The most common type is idiopathic scoliosis, which has no known singular underlying cause and patients are typically otherwise healthy.^18–20^ Idiopathic scoliosis is most frequently diagnosed in adolescence (adolescent idiopathic scoliosis, AIS) and is more prevalent in females than males.^18,19^ Scoliosis can also occur secondary to neuromuscular diseases (neuromuscular scoliosis, NMS) such as cerebral palsy, Duchenne muscular dystrophy, and spinal muscle atrophy, in which muscle weakness and spasticity impact spinal stability.^21^ The rapid progression of spinal deformity in NMS patients can lead to pain, skin ulceration, and cardiac and pulmonary dysfunction in addition to neuromuscular effects of the underlying disease.^21,22^ Scoliosis patients with severe curvatures (Cobb angle of >50%) may undergo spinal fusion surgery to stabilize and straighten the spine with the goal of preventing progression.^18–20^ Compared to AIS, NMS patients experience increased intra- and post-operative complications that affect neurological and pulmonary systems.^23^ They may also experience increased intra-operative blood loss, are more susceptible to wound infection and implant failures, and take longer to recover.^21,22^

Given the underlying conditions and surgical complications facing individuals with NMS, it was hypothesized that these patients would exhibit differences in wound inflammation. To address this, the cellular and cytokine content was measured in early post-operative wound drainage collected from AIS and NMS patients following spinal fusion surgery. NMS patients exhibited differences in the wound inflammatory response compared to AIS patients, which may impact the known healing issues with this patient population.

## Methods

### Ethics statement for human subjects studies

Studies performed using samples derived from human subjects obtained ethical approval from the Lifespan Institutional Review Board (IRB #1135267). Studies were performed in compliance with the 1975 Declaration of Helsinki.

### Spinal fusion surgery and drain placement

Spinal fusion surgery was performed on AIS or NMS patients. Wound drains were placed following surgery as part of the standard protocol for pediatric spinal fusions. For AIS patients, Hemovac Drains (Zimmer Biomet, Braintree, MA) were placed subcutaneously with removal typically occurring by the second or third postoperative day. For NMS patients, Jackson Pratt Drains (Cardinal Health, Dublin, OH) were placed in the submuscular space and typically left in place for the duration of hospitalization due to increased drainage output. NMS patients also typically underwent a multi-layered closure with muscle flaps to mitigate known healing issues with this group.

### Wound drainage collection

Daily surgical wound drainage samples were collected for the duration of fluid output. Samples collected on the same day as surgery were considered post-operative day (POD) 0. Samples were immediately frozen at -80°C for later use in cytokine assays. Additionally, POD 1 wound drainage samples were collected from a subset of patients and kept on ice for same-day cell isolation and flow cytometry analysis.

### Cytokine measurement

A multiplex bead-based cytokine assay (LEGENDPlex, BioLegend, San Diego, CA) was used to measure the concentration of 13 analytes in wound drainage samples from all patients: IL-1≤, IFN-*α*2, IFN-*γ*, TNF-*α*, CCL2, IL-6, CXCL8, IL-10, IL-12p70, IL-17A, IL-18, IL-23, and IL-33. The assays were performed following the manufacturer’s protocol. All samples underwent a single freeze-thaw prior to analysis. 2-fold and 50-fold dilutions of each sample were assayed in technical duplicate. Samples were acquired on an Attune NxT Flow Cytometer (ThermoFisher Scientific, Waltham, MA) and analysis was performed using BioLegend LEGENDPlex software (Qognit, San Calos, CA). IL-17A and IL-33 data were ultimately excluded because ≥40% of technical duplicate samples fell below the limit of detection of the assay.

### Wound drainage cell isolation

Fresh POD 1 samples were obtained from 6 AIS and 6 NMS patients for cell isolation. Samples were kept on ice for same-day processing. Samples were centrifuged at 250 x g at 4°C for 5 minutes and the supernatant was removed. Cell pellets were resuspended in 5 volumes of NH_4_Cl and incubated for 10 minutes at room temperature for red blood cell lysis. The samples were then centrifuged at 250 x g for 5 minutes and the supernatant was discarded. The NH_4_Cl lysis step was repeated once. Following centrifugation, the samples were resuspended in HBSS (no calcium, no magnesium, no phenol red, ThermoFisher Scientific, Waltham, MA) and a cell count was performed using a Moxi Z counter (Orflo, El Cajon, CA).

### Flow cytometry analysis

Flow cytometry analysis was performed on freshly isolated POD 1 samples from AIS and NMS patients to identify immune cell subsets. 1x PBS containing 0.1% BSA was used as an antibody diluent. All antibodies were from BioLegend (San Diego, CA). Samples were treated with Human TruStain FcX to block Fc receptors. Samples were then treated with the following anti-human antibodies in the presence of Fc block: CD45-FITC (Clone 2D1), CD56-APC (Clone QA17A16), CD14-APC-Fire750 (Clone 63D3), CD3-BV605 (Clone SK7), CD16-PE-Dazzle594 (Clone B73.1), and CD66b-PE-Cy7 (Clone QA17A51). Cells were then treated with fixable viability dye-eFluor506 (ThermoFisher Scientific, Waltham, MA), followed by fixation with 1% paraformaldehyde. Samples were acquired on an Attune NxT flow cytometer (ThermoFisher Scientific, Waltham, MA). Analysis was performed using FlowJo Software (BD Biosciences, Franklin Lakes, NJ).

### Statistics

Statistical analyses were performed in GraphPad Prism (Version 10.4.1). Categorical demographic data was analyzed using Fisher’s exact test and numerical demographic data was analyzed using the Mann Whitney test to determine differences between AIS and NMS groups. For cytokine time course data, a mixed effects model was used because sampling time points were absent for certain patients. Statistical significance between AIS and NMS groups at each time point was determined using Šidák’s multiple comparisons test. For flow cytometric data, the Mann-Whitney test was performed to determine statistical differences between AIS and NMS groups.

## Results

### Scoliosis patient cohort

For wound cytokine analysis, drainage samples were obtained from 68 patients: 54 with AIS and 14 with NMS. Samples were collected from a subset of these patients for cell isolation and analysis. All AIS and NMS patients were adolescent, with average ages of 14.5 and 16, respectively. AIS patients skewed female, as expected,^19^ while NMS patients did not. More NMS patients had asthma and developmental delay compared to AIS patients. The intra-operative time and volume of administered packed red blood cells (pRBCs) were higher, on average, for NMS patients compared to AIS. Post-operatively, 2 NMS patients were re-admitted with wound infection, in line with known susceptibility to infection.^21,23^ No AIS patients were re-admitted with wound infection (Table 1). Additionally, one NMS patient died several days post-operatively.

**Table 1.**
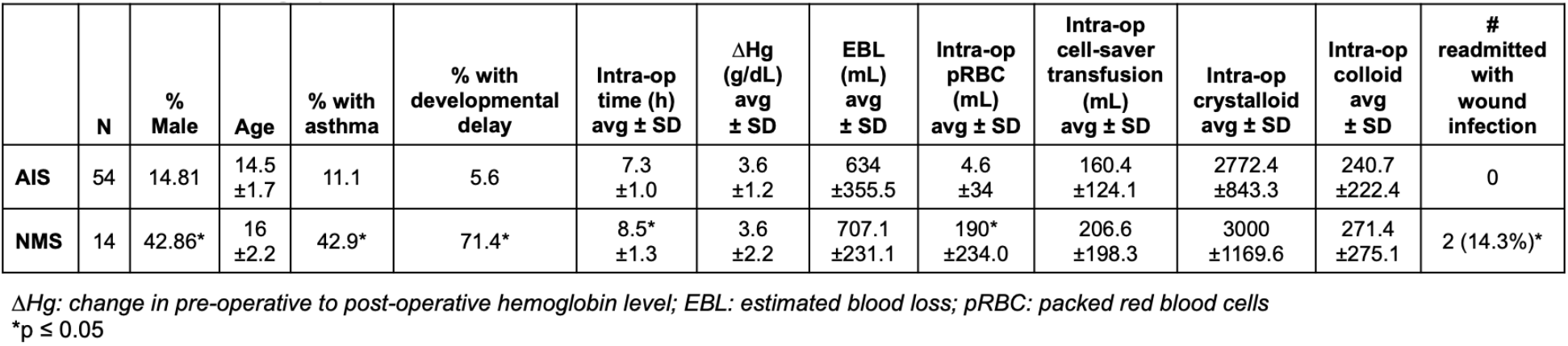
Patient demographic data.

### Cytokine kinetics in AIS and NMS wounds

NMS patients have associated conditions that can result in wound healing deficiencies, which we hypothesized would be reflected in an altered wound site inflammatory environment compared to AIS patients. To assess this, we screened wound drainage samples collected daily from POD 0 to POD 3 for a panel of pro-inflammatory cytokines with known roles in injury.^24–30^ Of the 13 cytokines assayed, 11 were sufficiently detected for inclusion in the analysis. In both AIS and NMS groups, several cytokines increased in abundance from POD 0 to POD 1 (IL-8, IL-6, CCL2, IL-1≤, TNF-*α*, IL-10, IFN-*α*2, and IL-12p70) (Figure 1A), while others showed minimal fluctuation across the timepoints sampled (IL-18, IL-23, IFN-*γ*) (Figure 1B). When comparing AIS and NMS samples, TNF-*α* and IFN-*α*2 were significantly elevated in the NMS group at POD 1. IL-8 and IL-1≤ had a similar trend of elevated POD 1 concentration in NMS samples, although the difference did not reach statistical significance. The remaining cytokines were detected at similar levels in both groups across all time points (Figure 1). These data demonstrate that NMS wounds are more inflamed than AIS wounds, although interestingly, only a subset of cytokines were elevated rather than a global elevation.

**Figure 1.**
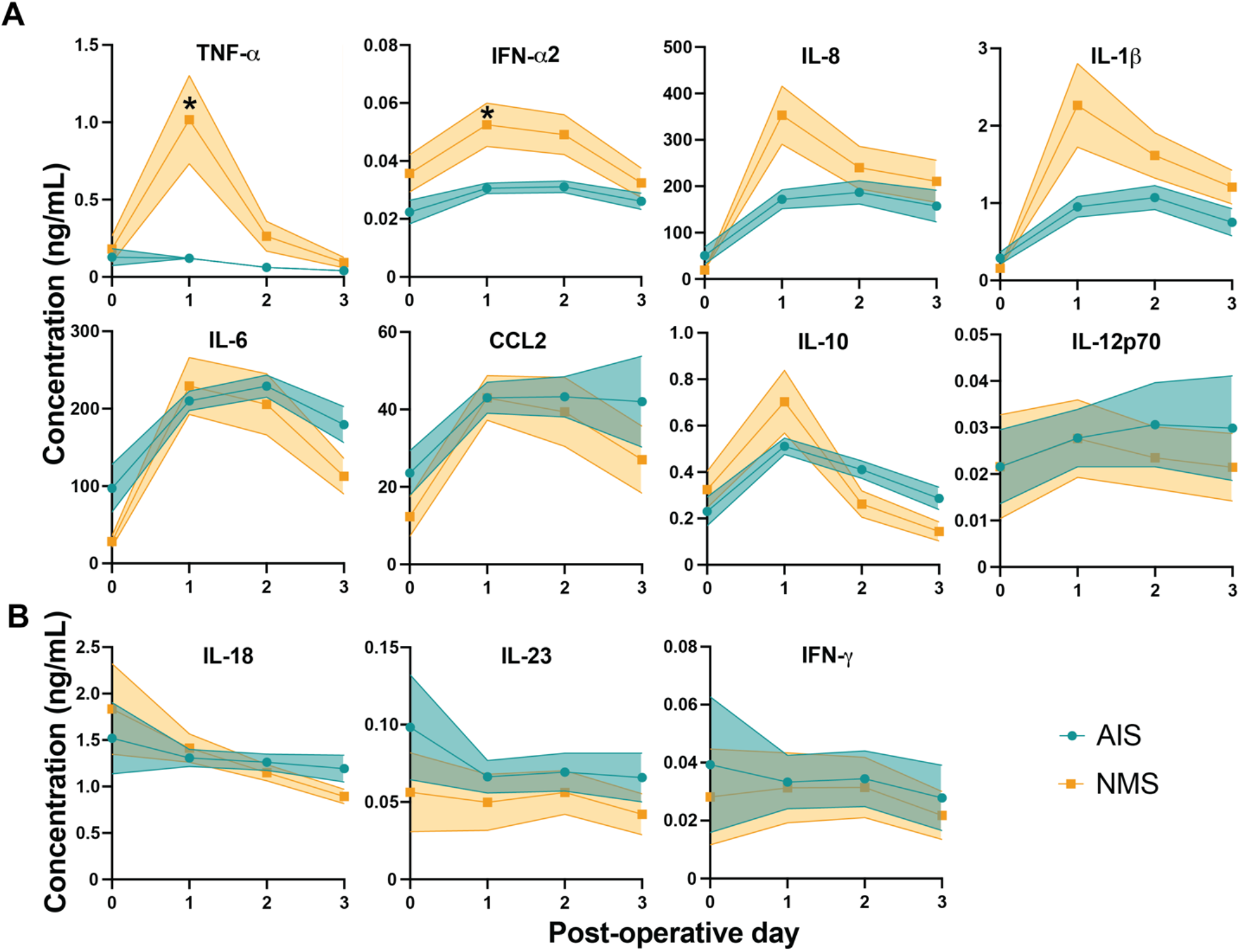
A comparison of cytokine concentrations between AIS and NMS surgical wound drainage samples revealed a subset of elevated wound-associated cytokines in NMS patient samples. (A) Several cytokines were elevated over time compared to post-operative day 0, while (B) others did not change over the assessed time course. Data shown are the average ± SEM, POD 0: AIS n = 11 and NMS n = 5; POD 1: AIS n = 50 and NMS n = 14; POD 2: AIS n = 45 and NMS n = 12; POD 3: AIS n = 24 and NMS n = 12. *p ≤ 0.05.

### AIS and NMS wound-associated leukocytes

Fresh wound drainage was acquired from a subset of patients and was assessed for the presence of cells with the goal of comparing AIS and NMS wound-associated leukocytes. Flow cytometry was used to identify wound leukocyte populations (Figure 2A). Cells isolated from wound drainage were abundant and >98% viable (Figure 2B). Nearly all isolated cells were CD45+ leukocytes. Over 95% of CD45+ cells were CD66b+ granulocytes (Figure 2C and 2D). The CD66b- population was a mix of CD3+ T cells, CD56+ NK cells, as well as CD14+CD16-, CD14-CD16+, and CD14-CD16-leukocytes, each of which formed a very small fraction of the overall leukocyte mix (Figure 2D). The viability of the neutrophils was surprising, as they can undergo many forms of cell death in response to stimulation.^31^ When comparing AIS and NMS groups, no differences were found in the distribution of leukocyte populations in the wound drainage samples.

**Figure 2.**
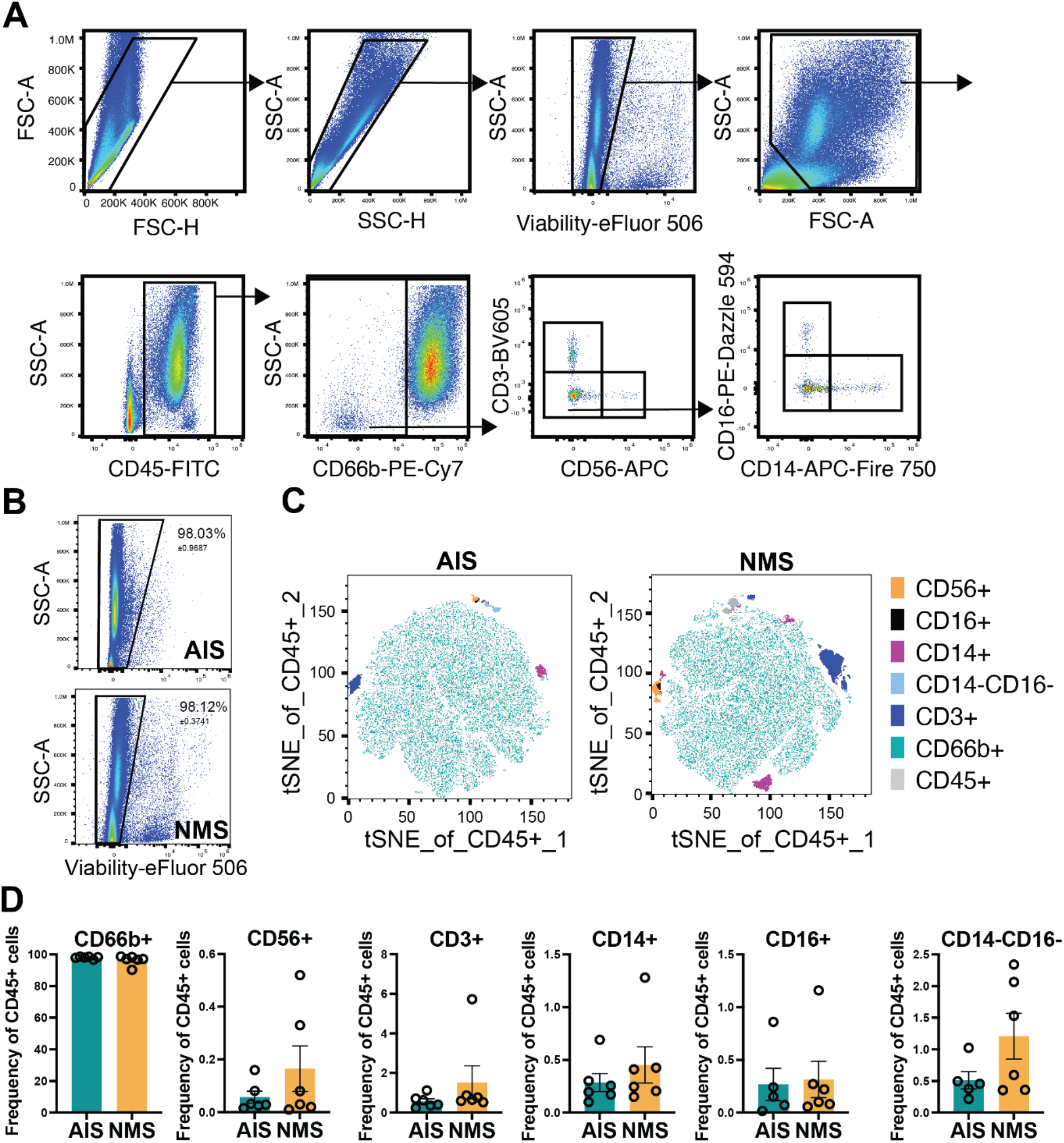
An immunophenotyping analysis was performed on surgical wound drainage samples from AIS and NMS patients. (A) Flow cytometry was used to identify CD45+ leukocyte subsets including CD66b+ granulocytes, CD3+ lymphocytes, CD56+ NK cells, as well as CD14+CD16-, CD14-CD16+, and CD14-CD16-leukocytes. (B) Cells isolated from surgical wound drainage were >98% viable. (C and D) CD66b+ granulocytes were the most abundant leukocyte population isolated from surgical wound drainage, as shown by t-SNE (C) and quantification of flow cytometry data (D). Data shown are the average ± SEM, n = 6 patients per group.

## Discussion

Surgical intervention is common to address the secondary health effects and quality of life issues that are associated with severe scoliosis. While most scoliosis patients fall into the idiopathic category with no associated health issues, individuals with underlying neuromuscular conditions face numerous health challenges that can affect their ability to heal from surgery. Compared to AIS patients, NMS patients experience longer post-operative healing times and are more susceptible to wound infection and other complications (Table 1),^23^ suggesting that wound healing responses may be impaired in these patients. It was hypothesized that NMS patients have an altered wound inflammatory response during the early acute stages of healing.

To address this hypothesis, we used spinal fusion surgical wound drainage to assess the wound-associated cellular and cytokine environment of AIS versus NMS patients at early post-operative time points. The data revealed that NMS wounds were more inflamed than AIS wounds, although the effect was targeted to specific cytokines, rather than a general trend of excess inflammation. Interestingly, IL-6, an important injury-associated cytokine, did not follow this trend.^28^ The two cytokines that had statistically significant increased expression in the NMS group, TNF-*α* and IFN-*α*2, are both essential to the wound healing response but can be pathogenic in excess. Many studies have shown that TNF*α* is elevated in diabetic and other chronic wound models, resulting in excessive fibroblast apoptosis, caspase activation, matrix-metalloprotease activity, and prolonged macrophage inflammation. TNF*α* blockade reversed these effects and allowed for progression to the proliferative and remodeling stages of healing, indicating its pathogenic role.^32–38^ Type I IFNs are less well characterized in the context of wound healing, although they are expressed in the injured tissue by plasmacytoid dendritic cells and keratinocytes and promote repair when present at normal levels. Studies in mice and humans have demonstrated that the exogenous addition of type I IFNs inhibits wound healing, suggesting that over-exuberant IFN-mediated inflammation can be detrimental to the progression of repair.^39,11,40,29,41,42^

Several factors could contribute to increased inflammation in NMS wounds, including differences in drain placement and longer intraoperative time, although this would likely cause a general increase in inflammatory markers, which we did not observe here. Immunological effects of the underlying neuromuscular conditions may also influence the cellular compositions of AIS and NMS wounds, which could influence cytokine profiles. To address this, we performed immunophenotyping of POD 1 wound drainage samples. Drainage samples were highly enriched for granulocytes, along with a very small proportion of lymphocytes, NK cells, and myeloid cells, although there were no compositional differences observed between the two groups. Of note, these results identify wound drainage as an ample source of viable neutrophils to facilitate functional studies. One limitation of this approach is that drainage may enrich for less adherent cells such that it is not entirely reflective of the wound cellular milieu. However, at this early post-operative time point, neutrophils are expected to be a dominant cellular player.^43,44,28,6^ While we could not link compositional changes to cytokine profiles in the AIS and NMS patients, this does not preclude functional differences of these cells and does not account for the role of cells absent from the drainage fluid. While these possibilities were outside of the scope of this study, they would be an important area of future work.

In sum, this study demonstrates that NMS and AIS patients have distinct cytokine profiles in draining surgical wound fluid, with a subset of inflammatory cytokines elevated in NMS patients in the early stages of wound healing compared to AIS patients. The overall cellular composition was similar, with neutrophils predominating in both patient groups. Furthermore, this study demonstrates the use of draining surgical wound fluid as a tool to analyze inflammatory responses in healing wounds, including both soluble factors and infiltrating immune cell subsets. This provides a framework for assessing wound-associated biomarkers from groups that are at risk of poor wound healing to better understand the repair response.

## Data Availability

All data produced in the present work are contained in the manuscript

## Authorship

MJC participated in study design, performed cellular and cytokine analyses, performed data analysis, and wrote the manuscript. RLM participated in study design, consented patients, and performed sample collection. ARDJ performed cellular analyses. CPE participated in study design, performed spinal fusion surgeries, and consented patients. AMJ participated in study design and wrote the manuscript.

## Conflict of Interest disclosure statement

The authors have no financial conflicts of interest to disclose.

## List of Abbreviations

AIS: adolescent idiopathic scoliosis
EBL: estimated blood loss
NMS: neuromuscular scoliosis
POD: post-operative day
pRBC: packed red blood cells

## Notes

### Competing Interest Statement

The authors have declared no competing interest.

### Funding Statement

This study did not receive any funding

